# Maternal Infection with Group B Streptococcus Correlates Inversely with Preeclampsia in Pregnant African American Women

**DOI:** 10.1101/2023.03.29.23287882

**Authors:** Keun Soo Kwon, Tzu Hsuan Cheng, Jordan Zhou, Huchong Cai, Sharon Lee, Ivan Velickovic, Mudar Dalloul, David Wlody, Ming Zhang

## Abstract

**Objective:** Preeclampsia is more prevalent in African American women than other races. GBS colonization rate in African American women is the highest among different races. We hypothesized there is an association between GBS colonization and preeclampsia among pregnant African American women.

**Study Design:** We evaluated the relationship between GBS colonization and preeclampsia in 11,766 patients’ records (93% were African Americans patients) between 2010 and 2017.

**Results:** The preeclampsia rate of GBS colonized patients (2.6%) was 59% less than that of GBS-negative patients (6.3%). This inverse correlation was independently significant after adjusting for other potential risk factors for preeclampsia: chronic hypertension, gestational diabetes mellitus and diabetes mellitus and other covariates. Further analysis among the patients without these risk factors also confirmed that the preeclampsia rate of GBS colonized patients (2.6%) was 55% less than that of GBS negative patients (5.7%). Similarly, patients with HSV+ or HIV+ had lower incidence of preeclampsia than those with HSV- or HIV-.

**Conclusions:** The unexpected finding of an inverse relationship between GBS colonization and preeclampsia suggests that the maternal immune response to GBS infection may be beneficial in reducing preeclampsia among African American patients.

## Introduction

Group B streptococcus (GBS) is a β-hemolytic, gram-positive bacterium that asymptomatically colonizes the lower genital tract (cervix, vagina and vulva) of approximately 18% of women worldwide [1, 2]. Globally, the prevalence of maternal GBS colonization is highest in Africa (∼22%) and lowest in Asia (∼11%) [3, 4]. In the United States, the estimated prevalence of maternal GBS colonization is about 19% [3]. During pregnancy, however, if GBS ascends from the vagina to the intrauterine space, it becomes highly pathogenic and is associated with numerous maternal and fetal complications such as preterm birth, stillbirth, and neonatal infections [1, 2]. In particular, GBS remains a leading cause of sepsis and meningitis in infants in the first 90 days of life [5]. Epidemiologic studies have also indicated that diabetes correlates with an increased risk for invasive GBS infections [6].

Reports that have focused on whether GBS colonization in pregnant women affects the incidence of preeclampsia are inconclusive. In a cross-sectional study using two statewide hospital databases (Florida, 2001: 190,645 pregnant women; Texas, 2004-2005: 577,153 pregnant women), the presence of GBS was inversely associated with pre-eclampsia. However, in a case-control study of 330 pregnant women at a hospital in Texas in 2010-2012, there was no association of GBS with preeclampsia [7]. More recently, a single center, retrospective cohort study of 60,029 births at Duke Health affiliated hospitals in North Carolina, during 2003-2015, also found that GBS colonization was not associated with preeclampsia [8].

The reasons for these discrepancies are not immediately apparent, but factors such as race and ethnicity may contribute. Preeclampsia is more prevalent in Black women than in women of other races [9, 10]. GBS colonization also varies among races, with Black women having the highest rate[3, 11-15]. In the cited studies, the percentage of patients who were Black ranged from 1.5%-35%. We hypothesized that if a predominantly Black population were studied, an association of GBS colonization and preeclampsia might be detected. As our hospital serves a predominantly African American community [16], we analyzed patient records from 2010 to 2017 to determine if there was such an association.

## Material and Methods

### Patients

Data from the hard copies of delivery records in Obstetric Department, along with electronic medical records, together which account for the record of every birth, were evaluated for all patients (mostly African American patients) who gave birth at University Hospital of SUNY Downstate between 2010 and 2017. Date of delivery, time of delivery, age and race of the mother, weeks of gestation at delivery, antepartum complications and mode of delivery, together with the corresponding clinical information for each patient, were compiled in a Microsoft Excel database. GBS swap was carried out by using eSwap transport system (COPAN Italia, Brescia, Italy) to swap the vagina and rectum. The sample was then transported to clinical pathology lab for GBS culture. The diagnosis and GBS’ sensitivity of penicillin were made by experienced clinical pathologist.

Statistical analyses were performed using SPSS software version 20 (IBM, New York, NY). We investigated the following potential relationships: preeclampsia and race, preterm birth and race, preterm birth and preeclampsia. Of total 11,766 patients delivered during 2010-2017, 93% were African American. We further categorized patients into following subgroups: 1) non-preeclampsia; 2) mild preeclampsia; 3) moderate preeclampsia; 4) severe preeclampsia, 5) eclampsia/HELLP syndrome, and 6) Preterm birth. Gestational Diabetes Mellitus (GDM) was diagnosed by ACOG’s two step approach: 1) Glucose challenging test showed blood sugar level ≥ 135 mg/dL; 2) Glucose three hour test showed elevated glucose concentrations at two or more time points. Diabetes Mellitus (DM) was diagnosed with two-hour Glucose challenging test which showed blood sugar level ≥ 200 mg/dL. Chronic hypertension was defined as systolic blood pressure (BP) ≥140 mm Hg and/or diastolic BP≥90 mm Hg documented either before pregnancy or before the 20th week of gestation on at least 2 separate occasions at least 4 hours apart. Preterm birth was defined as the birth of a baby before 37 weeks.

### Statistical Analyses

All data were expressed as means ± standard errors of means. Chi-square tests (2-sided) were used to determine whether GBS colonization positivity and preeclampsia were significantly associated. Logistic regression was used to calculate odds ratios (ORs) and 95% confidence intervals (CIs) that were adjusted for chronic hypertension (CHTN), diabetes mellitus (DM), gestational diabetes mellitus (GDM) and other covariates. A value of P<0.05 was considered statistically significant.

### Ethical approval

The Institutional Review Board of SUNY Downstate Health Sciences University, University Hospital of Brooklyn Downstate, approved this retrospective chart review (IRB approval# 973453-2).

## Results

The demographic information for 11,766 mostly African American pregnant women is summarized in Table 1. The rate of preeclampsia among these women was 5.8% (685 of 11,766 patients). This is much higher than the national average of 3.4% for all ethnic groups in the US [9, 17].

**Table 1.** Demographics of Patients and Antepartum Complication.

|  | Non-Preeclampsia | Preeclampsia | P-value * |
| --- | --- | --- | --- |
| Numbers of patients | 11,081 (94.2%) | 685 (5.8%) |  |
| Mean gestational age (weeks) | 36.7 ± 4.8 | 35.9 ± 3.7 | **0.000\*** |
| Mean age of patients (years) | 28.4 ± 6.4 | 27.9 ± 6.7 | 0.102 |
| BMI | 31.8 ± 7.2 | 32.5 ± 6.9 | **0.018\*** |
| Parity | 1.1 ± 1.4 | 1.1 ± 1.3 | 0.416 |
| Non-Smoking | 7607 (94.0%) | 487 (6.0%) | 0.719 |
| Smoking | 304 (94.7%) | 17 (5.3%) |  |
| Natural Pregnancy | 11074 (94.1%) | 689 (5.9%) | 1.000 |
| IVF Pregnancy | 7 (100%) | 0 (0%) |  |
| IUGR- | 11023 (94.2%) | 679 (5.8%) | 0.273 |
| IUGR+ | 58 (90.6%) | 6 (9.4%) |  |
| No history of preeclampsia | 11074 (94.2%) | 685 (5.8%) | 1.000 |
| With history of preeclampsia | 7 (100%) | 0 (0%) |  |
| GBS- | 9634 (93.7%) | 646 (6.3%) | **0.000\*** |
| GBS+ | 1447 (97.4%) | 39 (2.6%) |  |
| HSV- | 10777 (94.1%) | 676 (5.9%) | **0.025\*** |
| HSV+ | 304 (97.1%) | 9 (2.9%) |  |
| HIV- | 10954 (94.1%) | 683 (5.9%) | **0.035\*** |
| HIV+ | 127 (98.4%) | 2 (1.6%) |  |
| GDM- | 10835 (94.6%) | 619 (5.4%) | **0.000\*** |
| GDM+ | 246 (78.8%) | 66 (21.2%) |  |
| DM- | 11072 (94.2%) | 684 (5.8%) | 0.453 |
| DM+ | 9 (90.0%) | 1 (10.0%) |  |
| CHTN- | 10766 (94.2%) | 657 (5.8%) | 0.073 |
| CHTN+ | 315 (91.8%) | 28 (8.2%) |  |
| Term births | 8880 (95.7%) | 401 (4.3%) | **0.000\*** |
| Preterm births | 2201 (88.6%) | 284 (11.4%) |  |
| Cesarean section delivery | 3119 (91.9%) | 274 (8.1%) | **0.000\*** |
| Non- Cesarean section delivery | 7962 (95.1%) | 411 (4.9%) |  |

\* P < 0.05 indicates statistical significance.

Of the 11,766 patients, 1,486 were diagnosed clinically with GBS colonization during pregnancy (12.6%). Among these GBS colonized patients, 39 developed preeclampsia (a rate of 2.6%), a 59% decrease than the preeclampsia rate of patients who were GBS negative (646/10,280 or 6.3%; P<0.01; Fig. 1a).

**Figure 1.**
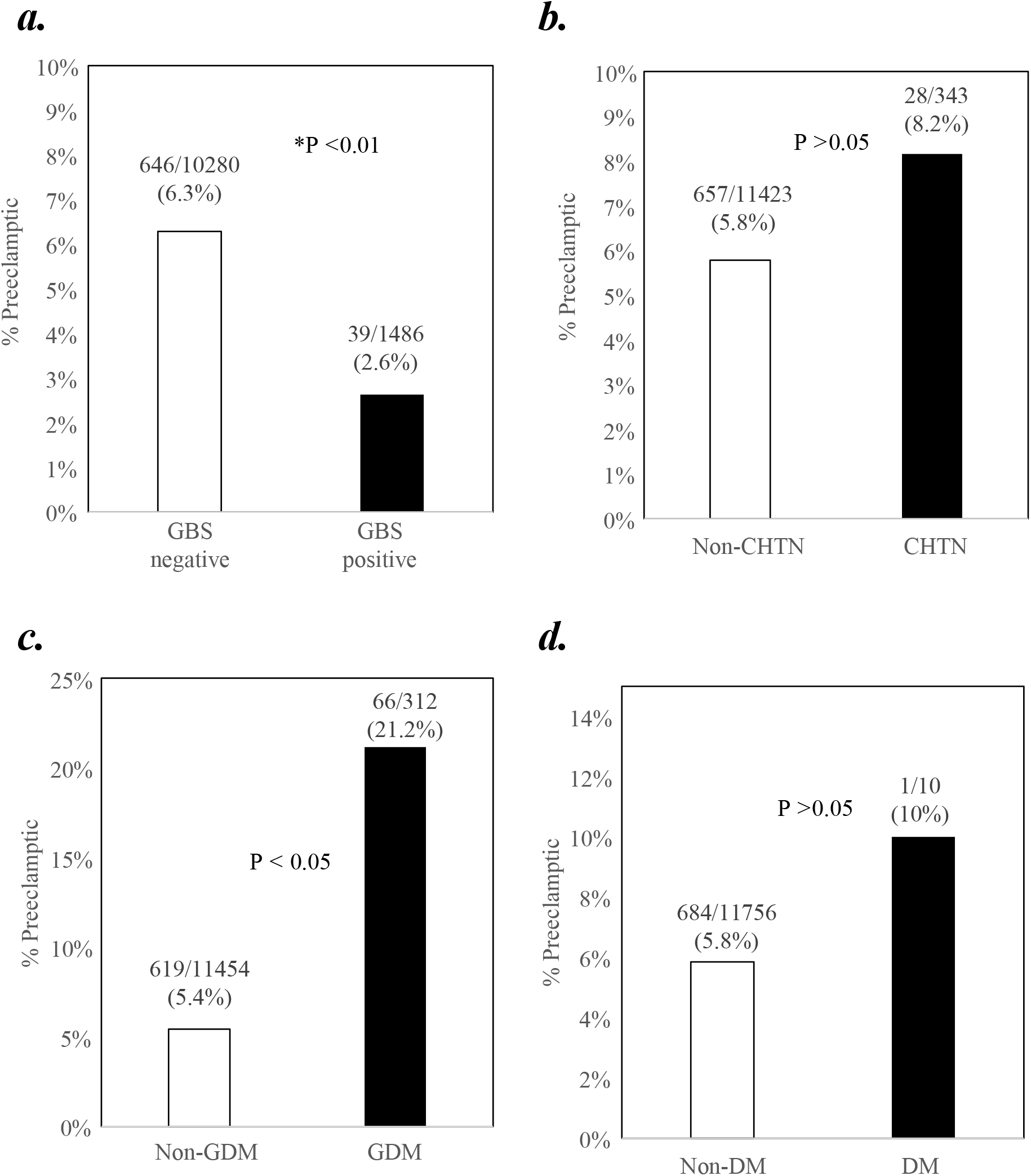
a) Comparison of the preeclampsia rates of GBS-negative and GBS-positive patients. b) Comparison of the preeclampsia rates of patients with and without CHTN. c) Comparison of the preeclampsia rates of patients with and without GDM. d) Comparison of the preeclampsia rates of patients with and without DM. * indicates statistical significance (P < 0.05).

CHTN, GDM and DM have been suggested to increase the risk of preeclampsia [18-25]. We investigated whether there was a relationship between these potential antepartum risk factors and preeclampsia. Of the 343 patients with CHTN (2.9% of the total), 8.2% (28/343) developed preeclampsia. By comparison, 5.8% (657/11,423) of the patients who did not have CHTN developed preeclampsia. Statistical analysis did not show a significant difference between these groups (Fig. 1b).

There were 312 patients who developed GDM (2.7% of the total). Among these patients, 21.2% (66/312) developed preeclampsia. In comparison, 5.4% (619/11,454) of the patients who did not have GDM developed preeclampsia. Statistical analysis showed a significant difference between these two groups (Fig. 1c).

Of the 10 patients with DM (0.1% of the total), 10% (1/10) developed preeclampsia. In comparison, 5.8% (684/11,756) of patients among the non-DM group developed preeclampsia. Statistical analysis did not show a significant difference between these two groups (Fig. 1d). Thus, CHTN and DM did not increase the risk of developing preeclampsia in these African American patients.

The mean gestational age at delivery of the preeclamptic patients was lower than that of non-preeclamptic patients (35.9 ± 3.7 vs. 36.7 ± 4.8; P <0.001; Table 1). These preeclamptic patients also had higher mean BMI than non-preeclamptic patients (32.5 ± 6.9 vs. 31.8 ± 7.2; P <0.05; Table 1). Interestingly, HSV+ patients had lower incidence of preeclampsia than HSV-patients (2.9% vs. 5.9%, P < 0.05; Table 1). Similarly, patients who were HIV+ also had lower incidence of preeclampsia than those that were HIV-(1.6% vs. 5.9%, P < 0.05; Table 1). In addition, there were significantly more preterm births in preeclamptic patients than non-preeclamptic patients (11.4% vs. 4.3%; P <0.01; Table 1). As expected, preeclamptic patients were also more likely to have C-sections than non-preeclamptic patients (8.1% vs. 4.9%; P < 0.001; Table 1).

To determine if the significance of a lower rate of preeclampsia among GBS colonized patients was confounded by the presence of CHTN, GDM, DM and other antepartum complications, logistic regression analyses were carried out. After adjusting for these potential confounders, we found that the inverse correlation between GBS colonization and preeclampsia remained independently significant (adjusted odds ratio=0.475; 95% confidence interval (C.I.): 0.327-0.690; P<0.01; Table 2). The accuracy of the regression model was 94.0%.

**Table 2.** Logistical Regression Analysis of Confounder Variables.

|  |  | B | S.E. | Wald | df | Sig.<sup>b</sup> | Adjust Odds Ratio (AOR) | 95% C.I. for AOR |  |
| --- | --- | --- | --- | --- | --- | --- | --- | --- | --- |
|  |  |  |  |  |  |  |  | Lower | Upper |
| Step 1<sup>a</sup> | GBS | -.744 | .190 | 15.324 | 1 | **.000\*** | .475 | .327 | .690 |
|  | BMI | .011 | .006 | 2.878 | 1 | .090 | 1.011 | .998 | 1.023 |
|  | Smoking | -.142 | .264 | .290 | 1 | .590 | .868 | .517 | 1.456 |
|  | Parity | -.031 | .037 | .712 | 1 | .399 | .970 | .903 | 1.042 |
|  | IVF | -19.207 | 19108.895 | .000 | 1 | .999 | .000 | 0.000 |  |
|  | IUGR | .365 | .488 | .558 | 1 | .455 | 1.440 | .553 | 3.749 |
|  | History of Preeclampsia | -19.066 | 16760.298 | .000 | 1 | .999 | .000 | 0.000 |  |
|  | HSV | -1.240 | .512 | 5.864 | 1 | **.015\*** | .289 | .106 | .790 |
|  | HIV | -1.887 | 1.020 | 3.421 | 1 | .064 | .152 | .021 | 1.119 |
|  | GDM | 2.066 | .178 | 135.119 | 1 | **.000\*** | 7.897 | 5.574 | 11.189 |
|  | DM | -18.069 | 13355.086 | .000 | 1 | .999 | .000 | 0.000 |  |
|  | CHTN | -.400 | .305 | 1.727 | 1 | .189 | .670 | .369 | 1.218 |
|  | Age | -.025 | .008 | 11.035 | 1 | **.001\*** | .975 | .961 | .990 |

- a. Variable(s) entered on step 1: GBS, BMI, Smoking, Parity, IVF, IUGR, History of Preeclampsia, HSV, HIV, GDM, DM, CHTN, Age. - b. \*P < 0.05 indicates statistical significance.

Further analysis was carried out among the patients without the confounding risk factors of CHTN, GDM, or DM (11,160/11,766 or 94.5% of the total patients in Fig. 1). Of the 11,160 patients, 1,394 were diagnosed clinically with GBS colonization during pregnancy (12.5%). Among these GBS colonized patients, 36 developed preeclampsia (a rate of 2.6%), a 55% decrease than the preeclampsia rate of patients who were GBS negative (5.7%; P<0.01; Fig. 2).

**Figure 2.**
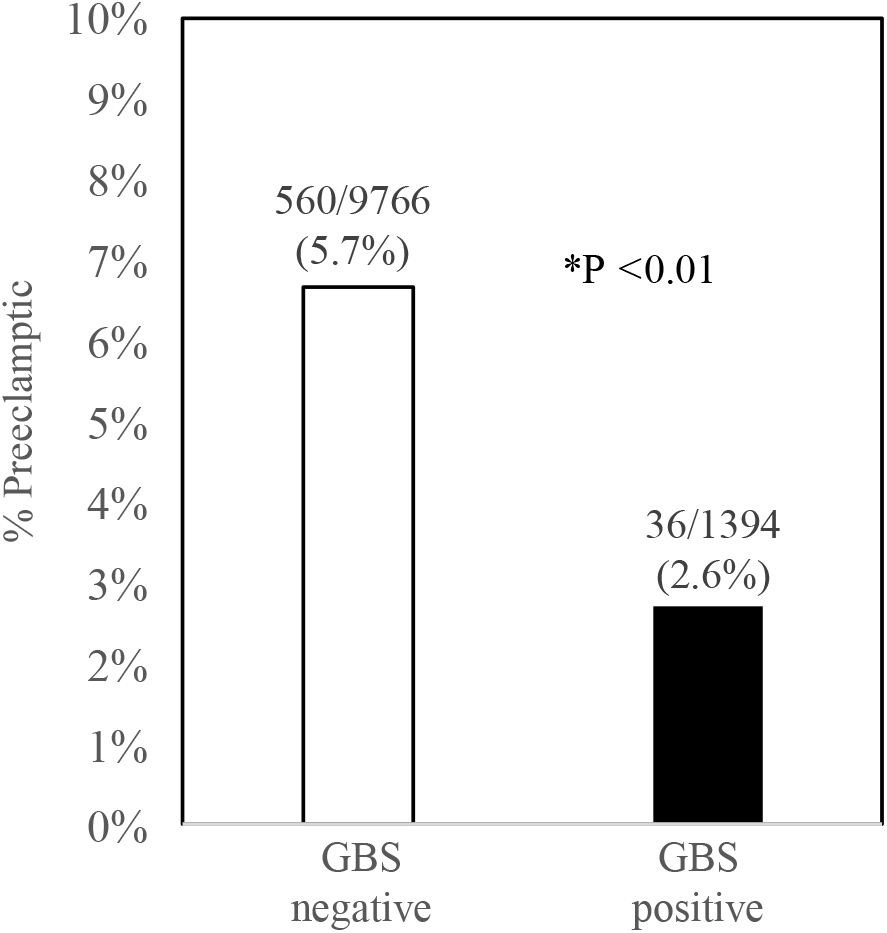
Comparison of the preeclampsia rates among the patients without the confounding risk factors of CHTN, GDM, or DM. * indicates statistical significance (P < 0.05).

We further analyzed the sub-types of preeclampsia and found that both moderate and severe sub-types of preeclampsia were significantly less among the GBS+ patients than GBS-patients (Fig. 3). There were similar trends of less moderate and severe sub-types of preeclampsia among the HSV+ patients compared to HSV-patients, but these were not statistically significant (Fig. 4).

**Figure 3.**
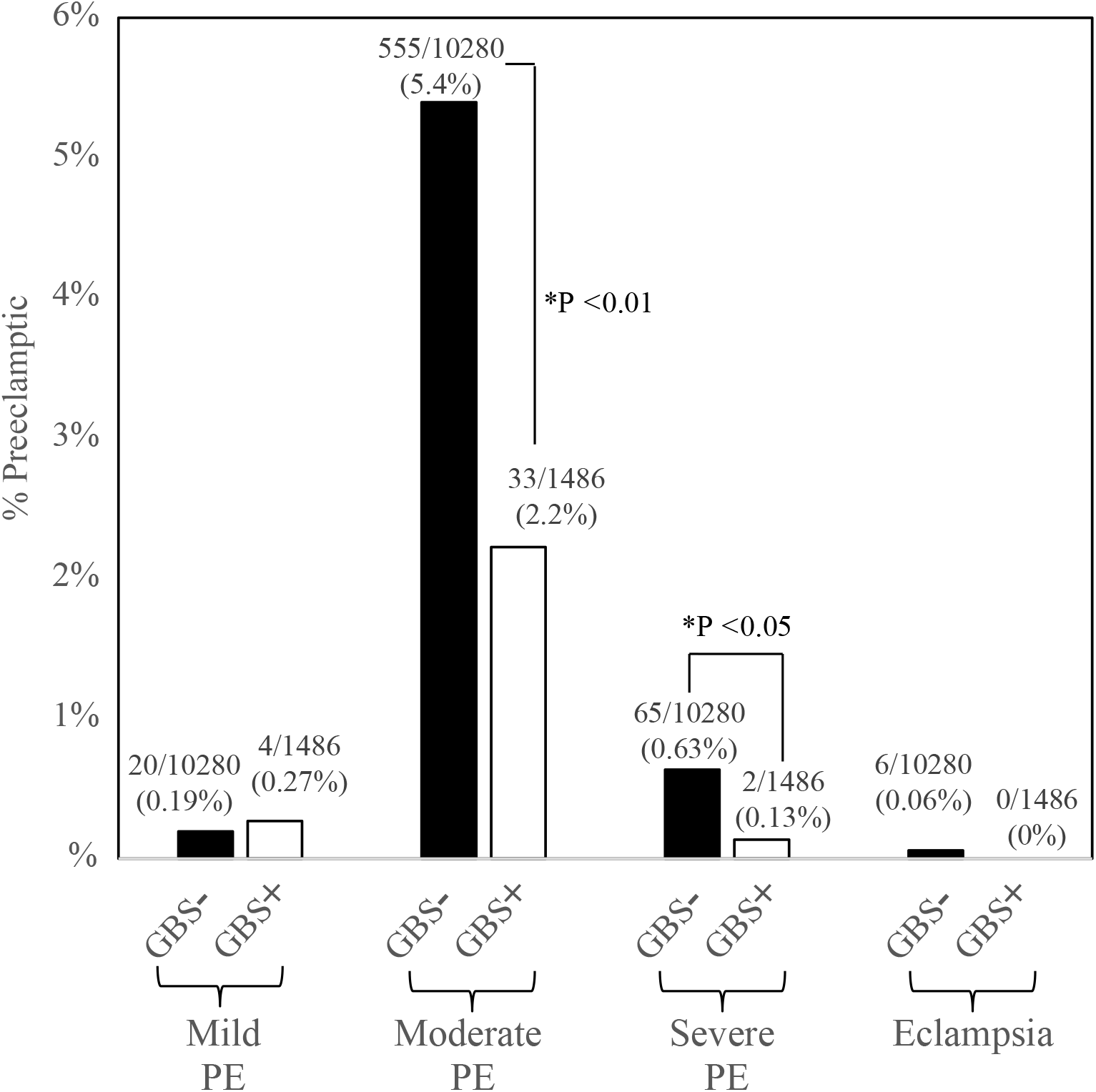
The rates of moderate and severe sub-types of preeclampsia were less in the GBS+ patients than those of GBS-patients. * indicates statistical significance (P < 0.05).

**Figure 4.**
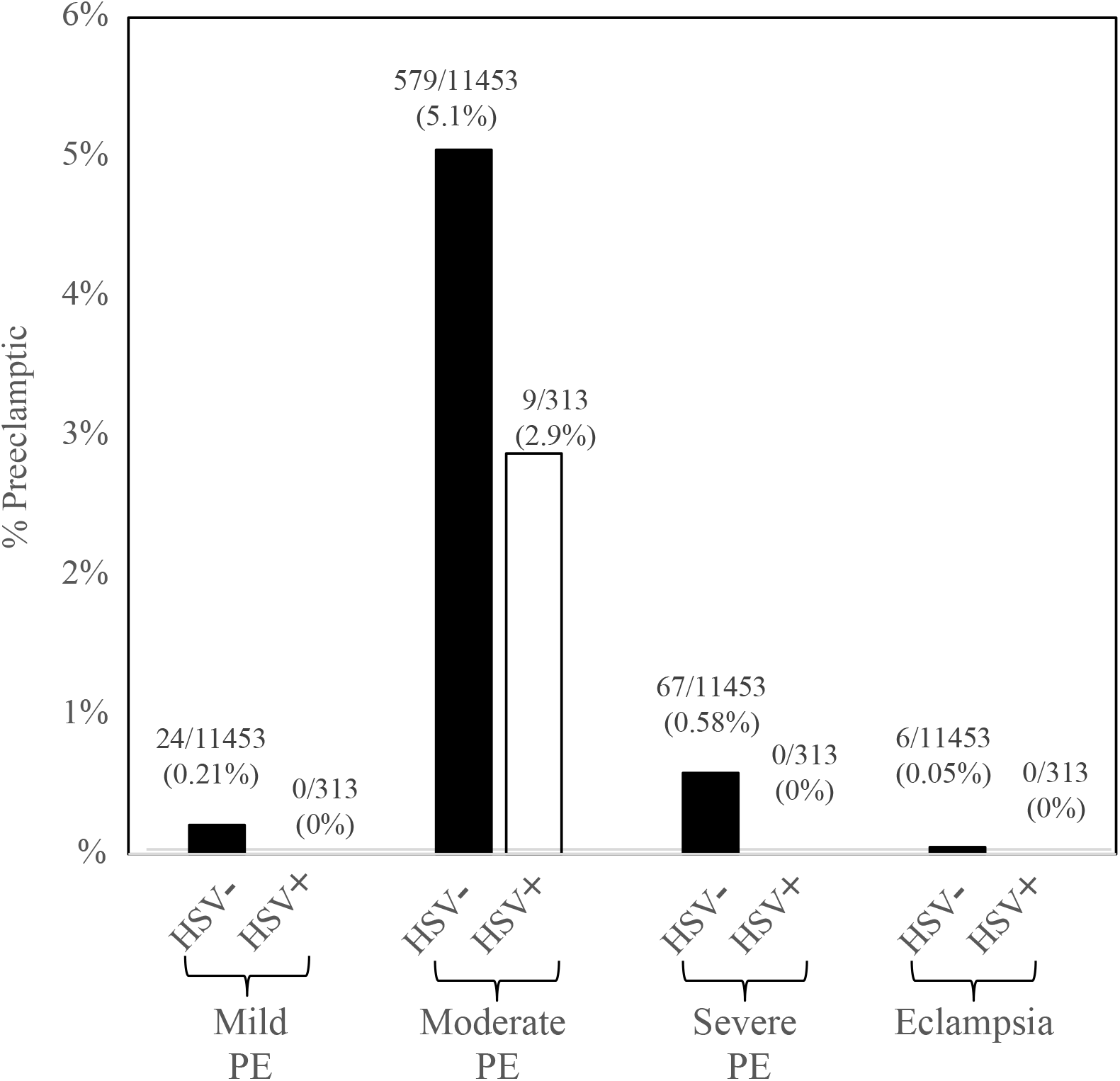
The rates of moderate and severe sub-types of preeclampsia show a trend of decrease in the HSV+ patients than those of HSV- patients, but did not reach statistical significance between sub-types.

## Discussion

This study showed that among pregnant African American women, GBS colonized patients had 59% decrease of the preeclampsia rate when compared with GBS negative patients (Fig. 1). This inverse correlation between GBS colonization and preeclampsia was independently significant after adjusting for the presence of CHTN, GDM, DM or other confounding risk factors (Table 2). Further analysis among the patients without the confounding risk factors of CHTN, GDM, or DM also confirmed that GBS colonized patients had 55% decrease of the preeclampsia rate when compared to GBS negative patients (Fig. 2).

The study reached the same conclusion found in the early 2000’s report using statewide hospital databases in Florida and Texas [26]. Interestingly, our study population had a much greater reduction (59%) than was found for the Florida (27%) and Texas (30%) cohorts. One possible explanation is that the denominators, the preeclampsia rates among the GBS negative patients, were lower in the Florida (3.8%) and Texas cohorts (4.2%) than in our study (6.7%), while the numerators, the preeclampsia rates for GBS colonized patients in Florida and Texas (2.8% and 2.9%), were similar to that in our study (2.7%). The higher rate of preeclampsia among our GBS negative patients compared with such patients in the Florida and Texas cohorts is not surprising as the patients in our study were mostly African Americans (93%), whereas African Americans comprised just 22% of the Florida cohort and 12% of the Texas cohort [26]. African American women are known to have a high risk of preeclampsia compared with other races [9]. Further studies are needed to confirm this inverse correlation between GBS infection and preeclampsia. If verified, the mechanism for such a reduction merits further investigation.

It has been speculated that immune regulation of the maternal-fetal interface is the result of the coordinated interaction between all its cellular components, including bacteria [27]. Accumulative evidence has indicated potential links of GBS infection with maternal/placental changes at cellular and molecular levels. Experimental studies showed that GBS inhibits autophagy in vaginal epithelial cells [28], causing vaginal epithelial exfoliation which facilitates spreading of infection to the uterus [29]. GBS also activates transcriptomic pathways related to premature birth in human extraplacental membranes [30]. GBS induces the expression of human *β*-defensin-2 in human extraplacental membrane [31], and secretion of maternal and placental cytokines, such as IL-1β, CXCL1, MMP-10 [32, 33], IL-1 receptor antagonist [34] and IL-10 [35]. Neutrophils respond to GBS by producing extracellular traps [36], which may be circumvented during GBS amniotic cavity invasion to induce fetal injury and preterm labor [37]. Human placental macrophages respond to GBS by releasing macrophage extracellular traps (METs) [38], possibly through activation of protein kinase D [39]. Decidual stromal cell-derived PGE2 regulates these macrophage responses [40]. Mast cells respond to GBS by degranulation, leading to the release of preformed and proinflammatory mediators [41], such as chymotrypsin-like cleavage specificity in response to GBS [42]. These maternal/placental changes by GBS at cellular and molecular levels may affect the development of preeclampsia.

Regarding the two previously cited case studies where GBS colonization was not correlated with preeclampsia, a likely explanation is the differences in race and geographic locations. In the case-control study of 330 patients at El Paso [7], only 1.5% of the patients were black (the rest were mostly Hispanic white), and only 8.5% of the total 330 of patients were GBS colonized (less than the 12.5% in our study; 12.3% in Florida cohort; 14.1% in Texas cohort [26]). The study of 60,029 patients at Duke Health affiliated hospitals^5^ contained 35% black patients, a GBS colonization rate of 21.6%, and a preeclampsia rate of 11.6%. There was no correlation between GBS colonization and incidence of preeclampsia. The overall GBS colonization and preeclampsia rates were much higher than those found in the present study and in the Florida/Texas cohorts study [26]. It is not clear if the 35% black patients, or the other race/ethnic groups present (44% white, 21% unknown) contributed to such high GBS colonization and preeclampsia rates.

Previous reports have suggested that CHTN, GDM, DM, BMI, parity, smoking, IVF pregnancy, intrauterine growth restriction (IUGR) and history of preeclampsia were all associated with an increased risk for preeclampsia [18-25]. In particular, nulliparity and pre-gestational diabetes have been suggested to predict preeclampsia [43]. We confirmed that GDM was positively correlated with preeclampsia (Fig. 1 and Table 2). While our study showed trends for higher rates of preeclampsia among African American patients with CHTN or DM, these trends were not statistically significant (Fig. 1). It is possible that with a larger sample size, we might obtain results showing associations with the statistical significance reported by others. Parity also did not independently predict preeclampsia in our cohort. One possible explanation may be that our patients were mostly African Americans, which differed from the patient populations of other studies.

Maternal GBS colonization is a major risk factor for perinatal infection, resulting in severe complications such as stillbirth and neonatal invasive disease. Considerable effort is being devoted to the development of GBS vaccines to minimize both maternal and fetal complications [44-46], with the most recent phase II clinical trial showing promise [47]. The unexpected finding of an inverse correlation between GBS infection and the development of preeclampsia in this study suggests that the maternal immune response to GBS infection is beneficial in reducing preeclampsia. In this scenario, the immune response to GBS would shift the focus of the immune system from targeting fetal antigens to producing anti-GBS antibodies, thus reducing the likelihood of preeclampsia. If verified, such an effect would add additional urgency to ongoing efforts to develop GBS vaccines to be made available for pregnant women to reduce maternal and fetal complications.

Like GBS, HSV+ patients had a lower preeclampsia rate than HSV-patients (Table 1). This inverse correlation between HSV and preeclampsia was independently significant after adjusting for other confounding risk factors (Table 2). Similarly, HIV+ patients also had a lower preeclampsia rate than HIV-patients (Table 1). However, such a correlation between HIV and preeclampsia was not significant after adjusting for other confounding risk factors (Table 2). Thus, at least in the population of our study, infection triggered immune responses seemed to alter the pathogenic process of preeclampsia. The underlying mechanism(s) is worthy of further investigation. Potential clinical applications of our findings may include vaccination of high risk pregnant patients to provide an additional benefit of preventing preeclampsia.

The concept of multi-markers screening for preeclampsia has been proposed by others. For example, a combination models using Elecsys data, second trimester uterine artery (UtA) Doppler ultrasonography measurements, and the serum fetoplacental protein levels used for Down’s syndrome screening, has been proposed in previous studies in predicting for preeclampsia [48]. Screening tools have also been proposed by National Institute for Health and Clinical Excellence (NICE), the American College of Obstetricians and Gynecologists (ACOG), and the Fetal Medicine Foundation (FMF). However, these tools currently may vary in their effectiveness due to the issues of health-care personnel education, economic coverage for biochemical markers and adjustment of algorithms based on population characteristics [49].

## Conclusions

Our study showed an inverse relationship between antepartum infections (e.g., GBS, HSV and HIV) and preeclampsia. The results suggest that the maternal immune response to infection may be beneficial in reducing preeclampsia among pregnant women.

## Data Availability

All data produced in the present work are contained in the manuscript

## Acknowledgements

The authors thank the helps of reviewing the charts and coordination by Philip Wan, Christopher Tenorio, Kenneth Ng, Joseph You, Joelle Hosenu, Maria Farag, Ian Lambert, and Joanna Serafin. We appreciate Dr. James Cottrell for his support and Dr. Julie Rushbrook for a critical reading of the manuscript.

